# Plasma biomarker profiles and the correlation with cognitive function across the clinical spectrum of Alzheimer’s disease

**DOI:** 10.1101/2021.02.05.21251241

**Authors:** Zhenxu Xiao, Xue Wu, Wanqing Wu, Jingwei Yi, Xiaoniu Liang, Saineng Ding, Li Zheng, Jianfeng Luo, Hongchen Gu, Qianhua Zhao, Hong Xu, Ding Ding

**Affiliations:** Institute of Neurology, Huashan Hospital, Fudan University, Shanghai, China; National Clinical Research Center for Aging and Medicine, Huashan Hospital, Fudan University, Shanghai, China; School of Biomedical Engineering, Shanghai Jiao Tong University, Shanghai, China; Department of Biostatistics, School of Public Health, Fudan University, Shanghai, China; Key Lab of Public Health Safety of the Ministry of Education, Fudan University, Shanghai, China

**Keywords:** Plasma p-tau181, Simoa, cognitive domain, CDR, Alzheimer’s disease

## Abstract

**Background:** Plasma biomarkers showed a promising value in the disease diagnosis and management of Alzheimer’s disease (AD). However, profiles of the biomarkers and the association with cognitive domains along the spectrum of cognitive performance deterioration have seldom been reported.

**Methods:** We recruited 320 individuals with cognitive impairment and 131 cognitively normal participants from a memory clinic and a community cohort. Participants were classified into 6 groups based on their Clinical Dementia Rating (CDR) scores and clinical diagnosis of AD, amnestic mild cognitive impairment, and normal cognition (NC). Each participant was administered the neuropsychological tests assessing the global and domain-specific cognition. Plasma Aβ_1-40_, Aβ_1-42_, Aβ_1-42_/Aβ_1-40_, total tau (t-tau), neurofilament protein light chain (NfL), and phosphorylated tau at threonine 181 (p-tau181) were quantified using the Single molecule array platform.

**Results:** Along with plasma Aβ_1-40_, Aβ_1-42_, Aβ_1-42_/Aβ_1-40_, t-tau, and NfL, p-tau181 significantly increased across the groups with the incremental CDR scores from NC (CDR = 0) to severe AD (CDR = 3). Compared with other biomarkers, p-tau181 had a stronger correlation with Global cognition (*r* = −0.494, *P* < 0.001), Memory (*r* = −0.417, *P* < 0.001), Attention (*r* = −0.388, *P* < 0.001), Visuospatial function (*r* = −0.328, *P* < 0.001), and Language (*r* = −0.123, *P* = 0.014). Among AD participants with CDR ≥ 1, higher p-tau181 was correlated with worse Global cognition (*r* = −0.295, *P* < 0.001), Memory (*r* = −0.172, *P* = 0.045), and Attention (*r* = −0.184, *P* = 0.031).

**Conclusions:** Plasma p-tau181 had a stronger correlation with cognitive domains than other biomarkers, especially in late-stage AD. It could reflect the AD pathology in vivo and may be a promising blood-based biomarker in clinical settings.

## Background

Alzheimer’s disease (AD), the most common cause of dementia, is characterized by the accumulation of the amyloid plaques and neuronal tangles in the brain [1]. Previous AD-relevant biomarkers could only be detected in the cerebrospinal fluid (CSF) or through positron emission tomography (PET) [1]. With the development of ultrasensitive immunoassays technique, detecting AD relevant biomarkers in blood samples became available [2]. Plasma biomarkers have a promising value in clinic usage due to the non-invasiveness, cost-effectiveness, and easy accessibility [3]. Following Aβ_1-42_, Aβ_1-40_, total tau (t-tau), neurofilament protein light chain (NfL), recently reported plasma phosphorylated tau at threonine 181 (p-tau181) showed better diagnostic performance and prognostic value in several cohort studies [3-9].

The cognitive performance is a pivotal indicator in AD management and efficacy evaluation. Previous studies found plasma Aβ_1-42_ [10], NfL [11, 12], and p-tau181 [3, 4, 13] were significantly different in participants with mild cognitive impairment (MCI) and AD compared with participants with normal cognition (NC). However, few studies depicted plasma biomarkers’ profiles based on the cognitive performance and compared their discrepancy during the entire course of AD. It would be valuable to depict the trajectory of different plasma AD biomarkers from NC until the severe cognitive impairment stage since the cognitive manifestations are the most concerned issues for both clinicians, patients, and the caregivers.

In addition, some studies investigated the relationships between plasma biomarkers and various cognitive domains [14-23]. However, they only focused on individual markers, and the results were incomparable or inconsistent due to the diverse testing platforms and study designs. It is also worthwhile to observe which biomarker has the best correlation with diverse cognitive domains.

In this study, we aimed to observe the trajectory of the plasma biomarkers profile, including Aβ_1-40_, Aβ_1-42_, Aβ_1-42_/Aβ_1-40_, t-tau, NfL, and p-tau181, across the clinical spectrum of AD evaluated with the clinical dementia rating (CDR) scale. We also intended to explore the correlation between these biomarkers and the domain-specific cognitive function, especially in different clinical AD stages.

## Methods

### Study participants

Participants with cognitive impairment were consecutively recruited from the memory clinic of the department of neurology, Huashan Hospital, Shanghai, China from November 2018 to September 2020. The inclusion criteria included: (1) consulted at the memory clinic due to memory complaints from herself (himself) or proxy; (2) able to cooperate with physical examinations and neuropsychological tests; (3) diagnosed with single-domain amnestic MCI (aMCI-s), multi-domain amnestic MCI (aMCI-m), or AD; (4) consent to the blood draw.

Participants in the Shanghai Aging Study (SAS) were eligible to be selected as the controls with NC. The SAS is a community-based longitudinal cohort in downtown Shanghai, China. The original purpose of SAS was to investigate the prevalence, incidence and risk factors for dementia and MCI among older residents in an urban community. The detailed study design and recruitment procedure have been published elsewhere [24]. In this study, the participants were selected from the third wave of follow-up between Jun and Oct in 2020 if they were: (1) 60 years or older; (2) able to cooperate with physical examinations and neuropsychological tests; (3) diagnosed with NC; (4) consent to the blood draw.

### Demographics and assessment of covariables

The demographic and lifestyle characteristics were acquired from the participants and/or proxy through a questionnaire. The educational background was defined as the years of formal education. Participants who smoked daily within the past month were regarded as cigarette smoking, and participants who had one serving of alcohol weekly during the past year were defined as alcohol consumption [25]. Hypertension and diabetes mellitus were confirmed by the medical records [24].

Apolipoprotein E (APOE) genotype was assessed by the Taqman single nucleotide polymorphism method using the blood or saliva samples collected from the participants [26]. The presence of at least one APOE ε4 allele was regarded as APOE ε4 positive.

### Neuropsychological tests

Comprehensive neuropsychological tests were administered by the certified psychometrists. All the tests were translated and adapted from western countries harmonized to Chinese culture, and were validated in Chinese population. Each participant from the memory clinic received a battery of neuropsychological tests, including: (1) Mini-Mental Status Examination (MMSE) [27]; (2) Montreal Cognitive Assessment-Basic (MoCA-B) [28-30]; (3) Auditory Verbal Learning Test [31, 32]; (4) Symbol Digit Modalities Test [33, 34]; (5) Rey-Osterrieth Complex Figure test [34]; (6) Boston Naming Test [35]; (7) Trail Making Test (TMT) [36]. For those who refused or were unable to complete the whole battery of tests, only MMSE and MoCA-B were administered. As for the participants with NC, a battery of similar neuropsychological tests was administered [31]: (1) MMSE [27]; (2) Auditory Verbal Learning Test [31, 32]; (3) Conflicting Instructions Task (Go/No-Go Task) [31]; (4) Stick Test [31]; (5) Modified Common Objects Sorting Test [31]; (6) TMT [36].

We extracted raw scores from above-mentioned neuropsychological tests to evaluate five clinically relevant cognitive domains including Memory, Attention, Visuospatial function, Language, and Executive function (Supplementary Table 1). *Z*-scores (*Z* = (raw score - mean)/standard deviation) of domains were computerized for analysis.

### Cognitive impairment severity and consensus diagnosis

All participants and their proxy were interviewed by two neurologists specialized in neurodegenerative diseases. The CDR, a semi-structured inventory, was used to assess the severity of cognitive impairment. It covered six cognitive, behavior, and function aspects, including memory, orientation, judgment and problem solving, community affairs, home and hobbies performance, and personal care. The neurologists scored on each aspect, taking into consideration information collected from both participants and proxy. The global CDR score was calculated automatically on biostat.wustl.edu/∼adrc/cdrpgm/index.html by inputting in each CDR box score, based on the Washington University CDR-assignment algorithm with a 0-3 scale index [37, 38].

Neurologists and neuropsychologists reached a consensus diagnosis of the cognitive function of each participant. The presence or absence of dementia was defined using the DSM-[criteria [39]. AD was diagnosed according to the NINCDS-ADRDA criteria [40]. The diagnosis of MCI was based on Petersen’s criteria [41]: (1) cognitive complaint by the subject, informant, nurse, or physician, with CDR = 0.5; (2) objective impairment in at least 1 cognitive domain. (3) essentially normal functional activities (determined from the CDR and the Activities of Daily Living evaluation); and (4) absence of dementia [42]. Because aMCI is more likely to progress to AD [43], we only included participants with 2 types of aMCI: (1) aMCI-s, memory impairment was required with no deficit in other domains; (2) aMCI-m, memory impairment plus at least1 additional deficit in another domain. NC participants had no memory complaint and have been confirmed cognitively intact through detailed neuropsychological assessment. For participants from memory clinic, laboratory tests and MR scan were performed to rule out the secondary causes related to cognitive impairment.

In this study, the continuum of AD was described based on the combination of CDR and the cognitive diagnosis: NC (CDR = 0), aMCI-s (CDR = 0.5), aMCI-m (CDR = 0.5), mild AD (CDR = 1), moderate AD (CDR = 2), and severe AD (CDR = 3) [37, 38].

### Plasma biomarkers measurement

Blood samples were collected in Ethylene Diamine Tetraacetic Acid (EDTA) plasma tubes and centrifuged (1,000 rpm, 4 °C) for 15 min. After the centrifugation, plasma was transferred into 1.5ml Eppendorf tubes and stored at−80 °C refrigerators.

Plasma Aβ_1-40_, Aβ_1-42_, t-tau, NfL, and p-tau181 were quantified using an ultra-sensitive Single molecule array (Simoa) (Quanterix, MA, US) on the automated Simoa HD-X platform per manufacturer’s instruction. The multiplex Neurology 3-Plex A kits (Cat. No. 101995), NF-light assay (Cat. No. 103186), and p-tau181 Assay Kit V2 (Cat. No.103714) were purchased from Quanterix and used accordingly. Technicians who performed the assay were blinded to the clinical data.

### Statistical Analyses

Mean and standard deviation (SD) were used to describe normally distributed continuous variables, while the median and quartile 1 (Q1) to quartile 3 (Q3) were used to describe the skewed distributed continuous variables. For categorical variables, number (n) and frequencies (%) were employed. One-way Analyses of Variance (ANOVA) and Kruskal-Wallis tests were used for comparing continuous variables, and Pearson Chi-square and Fisher exact tests were used to compare the categorical variables. For the comparisons among multi-groups, ANOVA and post hoc Tukey multiple comparison tests were used for variables with equal variance, while Welch and post hoc Games-Howell tests were used for variables with unequal variance.

Boxplots and points were used to present the distributions of original values of six plasma biomarkers. Levene’s tests were used for testing the homogeneity of variances in six groups with different cognition status. Associations between domain-specific *Z* scores and log-transformed plasma biomarker indexes (due to non-normal distributions [3]) were examined using partial Pearson correlation analyses with the adjustment for confounding variables. The heatmap matrix was implemented to visualize the adjusted partial correlation coefficients *r* in all participants. Positive correlations (*r* > 0) were exhibited in red, and negative correlations (*r* < 0) were exhibited in blue in the heatmap figure. The same correlation analyses were applied to participants with CDR = 0, CDR = 0.5, and CDR ≥ 1, respectively.

Two-sided *P□*< *□*0.05 was considered statistically significant. Data were analyzed using IBM SPSS Statistics for Windows, version 25.0 (IBMCorp., Armonk, N.Y., USA) [44] and R (version 4.0.2) [45]. Box plots were produced using the package ggplot2 in R. The heatmap was visualized by GraphPad Prism version 8.0.0 for Windows, GraphPad Software, San Diego, California USA (www.graphpad.com).

## Results

### Characteristics of the study participants

We recruited 451 participants, including 320 participants from the memory clinic cohort and 131 NC participants from the community cohort. The characteristics of study participants were shown in Table 1. Significant difference was found in gender (*P* = 0.011), age (*P* < 0.001), education years (*P* < 0.001), and APOE ε4 allele (*P* < 0.001) among six cognitive performance groups. The most severe cognitive-impairment group (CDR = 3) had the highest proportion of women (68.4 %), the lowest mean age (mean = 62.3), the shortest education years (mean = 5.6), and the largest proportion of positive APOE ε4 allele (57.9 %). We did not find the significant discrepancy in smoking, alcohol consumption, hypertension, and diabetes mellitus among six groups. All Neuropsychological tests’ scores were significantly different among six groups (all *P* < 0.001).

**Table 1.** Demographic characteristics and the neuropsychological assessments among study participants.

|  |  | Clinical diagnosis |  |  |  |  |  | *P value* |
| --- | --- | --- | --- | --- | --- | --- | --- | --- |
|  |  | NC | aMCI-s | aMCI-m | Mild AD | Moderate AD | Severe AD |  |
|  |  | CDR = 0 | CDR = 0.5 | CDR = 0.5 | CDR = 1 | CDR = 2 | CDR = 3 |  |
| *Demographic characteristics* | Total<br>(N = 451) | (N = 131) | (N = 39) | (N = 113) | (N = 67) | (N = 63) | (N = 38) |  |
| Gender, female, n (%) | 260 (57.6) | 82 (62.6) | 14 (35.9) | 69 (61.1) | 31 (46.3) | 38 (60.3) | 26 (68.4) | 0.011 |
| Age, y, mean (SD) | 68.6 (8.8) | 69.2 (7.1) | 69.6 (9.3) | 70.0 (8.0) | 67.7 (9.7) | 69.1 (10.8) | 62.3 (8.2) | <0.001 |
| Education years, mean (SD) | 10.3 (4.1) | 11.8 (2.9) | 13.6 (3.6) | 10.2 (3.6) | 10.3 (3.6) | 8.4 (4.6) | 5.6 (4.0) | <0.001 |
| Smoking, n (%) | 79 (17.5) | 17 (13.0) | 5 (12.8) | 23 (20.4) | 15 (22.4) | 12 (19.0) | 7 (18.4) | 0.529 |
| Alcohol consumption, n (%) | 77 (17.1) | 19 (14.5) | 4 (10.3) | 21 (18.6) | 18 (26.9) | 12 (19.0) | 3 (7.9) | 0.139 |
| Hypertension, n (%) | 168 (37.3) | 53 (40.5) | 17 (43.6) | 44 (38.9) | 23 (34.3) | 22 (34.9) | 9 (23.7) | 0.461 |
| Diabetes mellitus, n (%) | 68 (15.1) | 19 (14.5) | 7 (17.9) | 19 (16.8) | 8 (11.9) | 10 (15.9) | 5 (13.2) | 0.933 |
| APOE ε4 positive, n (%) | 161 (35.7) | 13 (9.9) | 12 (30.8) | 48 (42.5) | 30 (44.8) | 36 (57.1) | 22 (57.9) | <0.001 |
| *Neuropsychological tests* |  |  |  |  |  |  |  |  |
| MMSE score, mean (SD) | 23.3 (6.7) | 29.0 (1.3) | 27.7 (1.7) | 25.8 (1.9) | 21.9 (1.4) | 15.7 (2.3) | 7.4 (2.6) | <0.001 |
| Memory*, median [Q1, Q3] | -0.07 [-1.18, 0.72] | 0.87 [0.56, 1.50] | 0.16 [-0.23, 0.72] | -0.23 [-0.71, 0.72] | -0.71 [-1.18, -0.23] | -1.18 [-1.18, -0.82] | -1.18 [-1.18, -1.18] | <0.001 |
| Execution function*, median [Q1, Q3] | 0.43 [-1.25, 0.43] | 0.43 [0.43, 0.43] | 0.43 [0.43, 1.49] | -0.41 [-1.25, 1.28] | 0.43 [-1.25, 0.43] | -1.25 [-1.25, -0.41] | -1.25 [-1.25, -1.25] | <0.001 |
| Attention*, median [Q1, Q3] | 0.43 [-0.39, 0.83] | 0.83 [0.67, 0.83] | 0.69 [0.40, 0.83] | 0.43 [-0.21, 0.83] | -0.36 [-0.78, 0.43] | -1.46 [-1.99, -0.39] | -2.01 [-2.42, -2.01] | <0.001 |
| Language*, median [Q1, Q3] | 0.58 [-0.27, 0.72] | 0.72 [0.58, 0.72] | 0.58 [0.16, 0.72] | -0.27 [-0.66, 0.72] | -0.27 [-1.16, 0.72] | -0.27 [-1.26, 0.72] | -1.26 [-2.25, -0.27] | <0.001 |
| Visuospatial function*, median [Q1, Q3] | 0.31 [-0.63, 0.93] | 0.93 [0.62, 0.93] | 0.76 [0.31, 0.93] | 0.15 [-0.32, 0.62] | -0.63 [-1.25, -0.005] | -1.25 [-1.87, -0.32] | -1.87 [-2.18, -1.02] | <0.001 |

Note: \* Z score transformed. NC, normal cognition; aMCI-s, amnestic mild cognitive impairment-single domain; aMCI-m, amnestic mild cognitive impairment-multiple domains; AD, Alzheimer's disease; CDR, Clinical Dementia Rating Scale; SD, standard deviation; Q1, quartile 1; Q3, quartile 3; APOE, apolipoprotein E; MMSE, mini-mental state examination.

### Plasma biomarkers across groups with different cognitive performance

As shown in Figure 1, plasma Aβ_1-40_, Aβ_1-42_, and Aβ_1-42_/Aβ_1-40_ ratio showed a descending trend, while plasma t-tau, NfL, and p-tau181 exhibited an increasing trend across groups with the increasing CDR scores in general. With regard to Aβ_1-40_, Aβ_1-42_ and NfL, we found significant differences between participants with NC (CDR = 0) and AD (CDR ≥ 1) (Figure 1 A, B and E). Aβ_1-42_/Aβ_1-40_, and t-tau showed differences only between participants with NC (CDR = 0) and severe AD (CDR = 3) (Figure 1 C and D). There was no significant discrepancy of Aβ_1-40_, Aβ_1-42_, Aβ_1-42_/Aβ_1-40_, t-tau, or NfL among participants with different severity level of AD (CDR = 1, 2, or 3). P-tau181 gradually increased across the AD cognitive continuum, with the lowest concentration in NC participants (CDR = 0), an increase in participants with aMCI (CDR = 0.5), and the highest concentration in participants with AD (CDR ≥ 1) (Figure 1 F). Specifically, participants with severe AD (CDR = 3) had significantly higher p-tau181 than those with mild AD (CDR =1).

**Figure 1.**
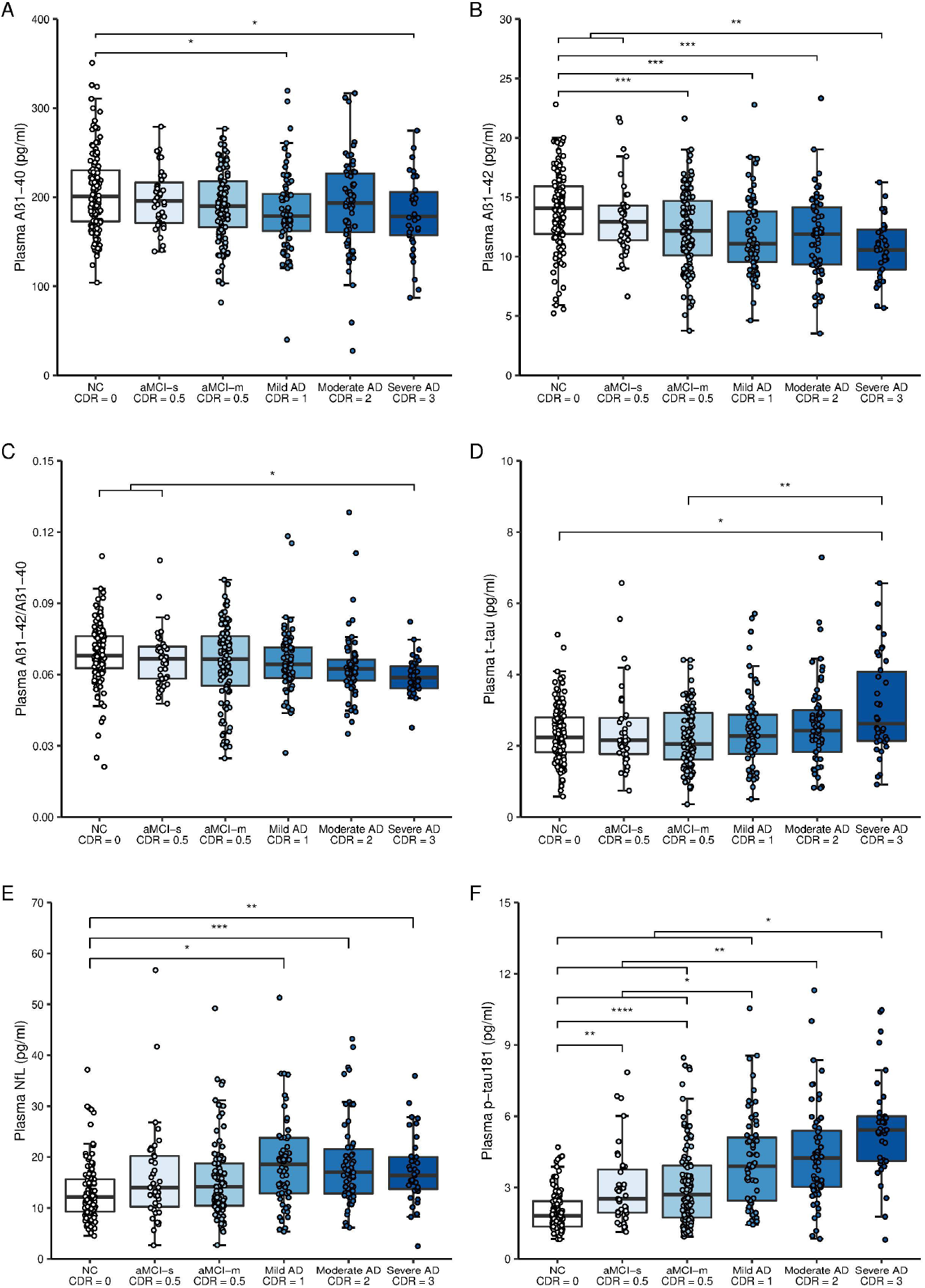
Plasma biomarkers in participants with different clinical cognitive status. Note: The ANOVA and the post hoc Tukey test were used for comparison of plasma Aβ_1-40_ & Aβ_1-42_ in six groups, while Welch test and the post hoc Games-Howell test were used to compare the plasma Aβ_1-42_/Aβ_1-40_, t-tau, NfL, p-tau181 among six groups. Six extreme values were not shown in panel E, but they were included in the statistical analyses. *P* values are presented with asterisks: ^*^ *P* < 0.05, ^* *^ *P* < 0.01, ^* * *^ *P* < 0.001, ^* * * *^ *P* < 0.0001. NC, normal cognition; aMCI-s, amnestic mild cognitive impairment-single domain; aMCI-m, amnestic mild cognitive impairment-multiple domains; AD, Alzheimer’s disease; CDR, Clinical Dementia Rating Scale; Aβ, amyloid-beta protein; t-tau, total tau; NfL, neurofilament protein light chain; p-tau181, tau phosphorylated at threonine 181.

### Correlation between plasma biomarkers and domain-specific cognition

Figure 2 showed the partial correlation matrix between six plasma biomarkers and six domain-specific cognitions after adjusting age, gender, education years, and APOE. Aβ_1-40_ was only correlated positively with Memory (*r* = 0.125, *P* = 0.012). Aβ_1-42_ was correlated positively with MMSE (*r* = 0.119, *P* = 0.012), Memory (*r* = 0.203, *P* < 0.001), and Attention (*r* = 0.104, *P* = 0.036), while Aβ_1-42_/Aβ_1-40_ was correlated positively with Memory (*r* = 0.114, *P* = 0.022) and Attention (*r* = 0.100, *P* = 0.044). T-tau had an inverse correlation with MMSE (*r* = −0.154, *P* = 0.001), Memory (*r* = −0.146, *P* = 0.003), and Attention (*r* = −0.116, *P* = 0.020). Higher NFL was correlated with worse MMSE (*r* = −0.298, *P* < 0.001), Memory (*r* = −0.263, *P* < 0.001), Attention (*r* = −0.209, *P* < 0.001), Visuospatial function (*r* = −0.236, *P* < 0.001), and Language (*r* = −0.105, *P* < 0.036). P-tau181 showed a stronger correlation inversely with MMSE (*r* = −0.497, *P* < 0.001), Memory (*r* = −0.417, *P* < 0.001), Attention (*r* = −0.388, *P* < 0.001), Visuospatial function (*r* = −0.328, *P* < 0.001), but a weaker correlation with Language (*r* = −0.123, *P* = 0.014).

**Figure 2.**
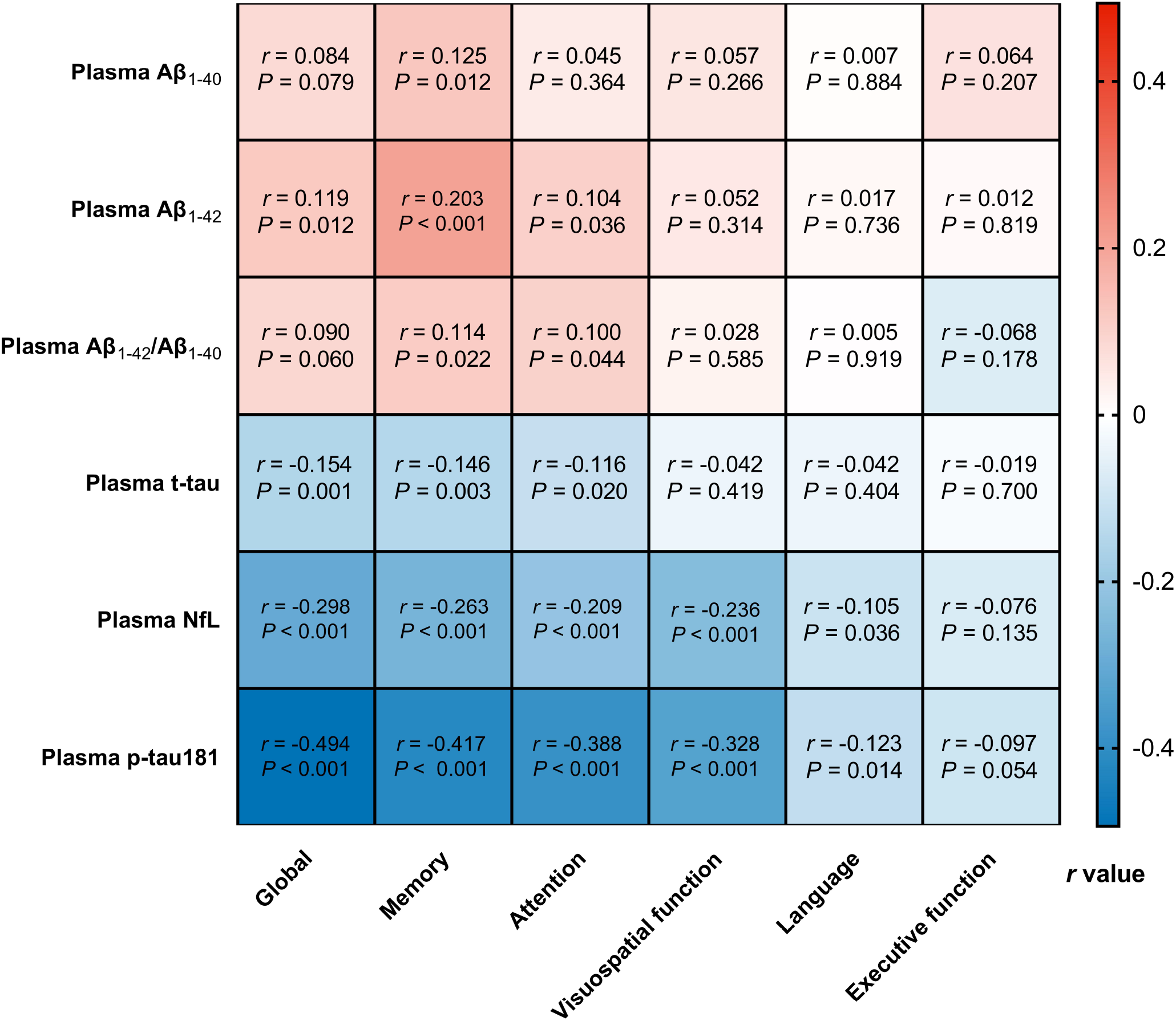
Correlations between plasma biomarkers and cognitive domains. Note: The plasma biomarkers concentrations were log transformed. The Pearson correlation coefficients (*r*) were adjusted for age, gender, education year, and APOE. Aβ, amyloid-beta protein; t-tau, total tau; NfL, neurofilament protein light chain; p-tau181, tau phosphorylated at threonine 181.

As shown in Table 2, no significant correlation was found between plasma biomarkers and cognitive domain scores in NC participants. Among aMCI participants with CDR = 0.5, higher Aβ_1-42_/Aβ_1-40_ was correlated with the deterioration of MMSE (*r* = −0.209, *P* = 0.011) and Executive function (*r* = −0.197, *P* = 0.021). Higher NfL was correlated with worse Visuospatial function (*r* = −0.174, *P* = 0.045), and higher p-tau181 was correlated with worse Memory (*r* = −0.167, *P* = 0.048).

**Table 2.** Correlations between plasma biomarkers and global & domain-specific cognition in participants with different AD stages.

| Biomarkers | Global (MMSE) |  |  |  |  |  | Memory |  |  |  |  |  | Attention |  |  |  |  |  |
| --- | --- | --- | --- | --- | --- | --- | --- | --- | --- | --- | --- | --- | --- | --- | --- | --- | --- | --- |
|  | CDR = 0 |  | CDR = 0.5 |  | CDR ≥ 1 |  | CDR = 0 |  | CDR = 0.5 |  | CDR ≥ 1 |  | CDR = 0 |  | CDR = 0.5 |  | CDR ≥ 1 |  |
|  | *r* | *P* | *r* | *P* | *r* | *P* | *r* | *P* | *r* | *P* | *r* | *P* | *r* | *P* | *r* | *P* | *r* | *P* |
| Plasma Aβ<sub>1-40</sub> | -0.105 | 0.256 | 0.097 | 0.240 | -0.032 | 0.681 | -0.088 | 0.350 | 0.048 | 0.574 | 0.083 | 0.331 | -0.094 | 0.311 | -0.052 | 0.542 | -0.060 | 0.480 |
| Plasma Aβ<sub>1-42</sub> | -0.086 | 0.350 | -0.109 | 0.188 | 0.067 | 0.394 | -0.023 | 0.805 | 0.148 | 0.080 | **0.184** | **0.030** | -0.169 | 0.068 | -0.020 | 0.814 | 0.038 | 0.659 |
| Plasma Aβ<sub>1-42</sub>/Aβ<sub>1-40</sub> | -0.023 | 0.804 | **-0.209** | **0.011** | **0.156** | **0.046** | 0.028 | 0.767 | 0.127 | 0.135 | 0.112 | 0.190 | -0.122 | 0.190 | -0.013 | 0.877 | **0.187** | **0.028** |
| Plasma t-tau | -0.008 | 0.933 | -0.108 | 0.193 | -0.132 | 0.092 | -0.182 | 0.051 | -0.151 | 0.074 | -0.063 | 0.463 | -0.119 | 0.203 | -0.079 | 0.353 | -0.042 | 0.622 |
| Plasma NfL | -0.168 | 0.067 | -0.052 | 0.530 | -0.129 | 0.104 | 0.048 | 0.610 | -0.083 | 0.330 | -0.158 | 0.067 | -0.053 | 0.568 | -0.026 | 0.762 | 0.030 | 0.732 |
| Plasma p-tau181 | -0.011 | 0.907 | -0.076 | 0.360 | **-0.295** | **<0.001** | -0.025 | 0.793 | **-0.167** | **0.048** | **-0.172** | **0.045** | 0.068 | 0.467 | 0.005 | 0.949 | **-0.184** | **0.031** |

| Biomarkers | Visuospatial function |  |  |  |  |  | Language |  |  |  |  |  | Executive function |  |  |  |  |  |
| --- | --- | --- | --- | --- | --- | --- | --- | --- | --- | --- | --- | --- | --- | --- | --- | --- | --- | --- |
|  | CDR = 0 |  | CDR = 0.5 |  | CDR ≥ 1 |  | CDR = 0 |  | CDR = 0.5 |  | CDR ≥ 1 |  | CDR = 0 |  | CDR = 0.5 |  | CDR ≥ 1 |  |
|  | *r* | *P* | *r* | *P* | *r* | *P* | *r* | *P* | *r* | *P* | *r* | *P* | *r* | *P* | *r* | *P* | *r* | *P* |
| Plasma Aβ<sub>1-40</sub> | -0.108 | 0.250 | -0.024 | 0.785 | 0.099 | 0.280 | -0.152 | 0.103 | -0.093 | 0.272 | -0.040 | 0.641 | -0.011 | 0.908 | 0.041 | 0.631 | -0.002 | 0.984 |
| Plasma Aβ<sub>1-42</sub> | -0.004 | 0.963 | -0.149 | 0.089 | 0.114 | 0.217 | -0.083 | 0.375 | -0.127 | 0.133 | -0.010 | 0.904 | -0.074 | 0.442 | -0.117 | 0.169 | 0.062 | 0.471 |
| Plasma Aβ<sub>1-42</sub>/Aβ<sub>1-40</sub> | 0.092 | 0.327 | -0.131 | 0.137 | 0.049 | 0.593 | 0.090 | 0.337 | -0.098 | 0.249 | 0.036 | 0.673 | -0.092 | 0.339 | **-0.197** | **0.021** | 0.085 | 0.322 |
| Plasma t-tau | 0.048 | 0.607 | 0.117 | 0.183 | -0.102 | 0.270 | -0.007 | 0.943 | -0.058 | 0.492 | -0.025 | 0.775 | 0.003 | 0.973 | 0.017 | 0.843 | -0.032 | 0.711 |
| Plasma NfL | -0.047 | 0.616 | **-0.174** | **0.045** | 0.042 | 0.652 | -0.147 | 0.115 | 0.038 | 0.654 | 0.007 | 0.932 | 0.056 | 0.558 | 0.127 | 0.136 | **-0.171** | **0.048** |
| Plasma p-tau181 | 0.079 | 0.398 | -0.044 | 0.616 | -0.066 | 0.479 | -0.025 | 0.793 | 0.071 | 0.405 | 0.067 | 0.438 | 0.006 | 0.948 | 0.133 | 0.120 | -0.060 | 0.489 |

Note: The plasma biomarkers concentrations were log transformed. The Pearson correlation coefficients (*r*) were adjusted for age, gender, education year, and APOE. Bold numbers indicate *P* < 0.05.
CDR, Clinical Dementia Rating Scale; Aβ, amyloid-beta protein; t-tau, total tau; NfL, neurofilament protein light chain; p-tau181, tau phosphorylated at threonine 181.

As for the AD participants with CDR ≥ 1, plasma Aβ_1-42_ had positive correlation only with Memory (*r* = 0.184, *P* = 0.030), and Aβ_1-42_/Aβ_1-40_ had positive correlation with MMSE (*r* = 0.156, *P* = 0.046) and Attention (*r* = 0.187, *P* = 0.028). Increased plasma NfL was only correlated with worse Executive function (*r* = −0.171, *P* = 0.048). Higher p-tau181 was correlated with worse MMSE (*r* = −0.295, *P* < 0.001), Memory (*r* = −0.172, *P* = 0.045), and Attention (*r* = −0.184, *P* = 0.031).

## Discussion

The present study demonstrated that plasma Aβ_1-40_, Aβ_1-42_, and Aβ_1-42_/Aβ_1-40_ had a decreasing trend, while plasma t-tau, NfL, and p-tau181 escalated along the deterioration of the cognitive performance. P-tau181 was the best indicator of clinical cognitive performance and had a stronger correlation with global and other cognitive domains than other five AD biomarkers. To our knowledge, it was the first study to exhibit the distribution of plasma p-Tau 181 using the Simoa HD-X platform in Chinese older individuals along the clinical AD continuum.

Pathology is pivotal in the diagnosis of AD. However, cognitive performance, including various cognitive domains, played a more significant role in patient management and efficacy evaluation. Cognitive manifestations are closely related to one’s daily function and quality of life for both patients and caregivers. Thus, we focused on individual’s performance, and classified the participants into six groups according to CDR levels and clinical cognitive diagnosis. One previous study found plasma p-tau181 increased at preclinical AD and further increased at the MCI and dementia stages [3]. Another group verified that plasma p-tau181 gradually increased from the Aβ-negative cognitively unimpaired older adults, through the Aβ-positive cognitively unimpaired and MCI groups, to the highest concentrations in Aβ-positive MCI and AD groups [4]. Our results testified their findings from the clinical perspective and further indicated that p-tau181 was a symptom-related plasma biomarker. Although some AD markers showed a significant escalating or descending trend along the whole disease continuum, plasma p-tau181 is the only one that had the significant discrepancy in the later stage of AD with overt dementia symptoms. This means plasma p-tau181 may keep increasing along with the deterioration of cognitive function, till the most severe stage of AD. Previous studies showed that plasma p-tau181 was correlated with CSF p-tau181 in AD patients, suggesting that plasma p-tau181 was originated from the central nervous system [3, 4]. Peripheral p-tau reflects the phosphorylation of the tau protein, which eventually leads to the neurofibrillary tangles in the brain [8]. Therefore, the continuous increment of the plasma p-tau across different stages of AD indicated the ongoing tau-related pathologic change in the brain, even in the late stage of the disease. This may potentially guide the development of disease-modifying therapy for terminal-stage patients in the future.

One subset cohort in a study demonstrated that plasma p-tau181 was correlated with baseline MMSE [4]. However, the relationships between plasma p-tau181 and different cognitive domains have not been reported. Our study revealed a strong correlation between plasma p-tau181 and AD-related cognitive domains. The correlations between p-tau181 and MMSE, Memory, and Attention were only observed in participants with dementia symptoms. This phenomenon may mainly reflect the fact that plasma p-tau181 is regulated differently by the disease staging, namely the Alzheimer’s pathological status in the brain. Another possibility is that, the narrow range of testing scale weakens the correlation of plasma biomarkers with cognition, due to “ceiling effects” or “floor effects” of the tests. However, this might not be the major explanation, because when analyzing with the more sensitive neuropsychological tests assessing Memory and Attention, the correlations were still not be observed in the subgroup with CDR = 0.

The NfL was an age-related biomarker [46]. Lin et. al. found that higher plasma NfL levels correlated with lower MMSE scores [15]. However, they did not adjust for age in the correlation analysis. The results from Chatterjee et. al. showed that plasma NfL was inversely correlated with working memory, executive function, and the global composite score after considering the age [14]. Two studies found plasma NfL associated with all cognitive domains after adjusting potential confounders including age [22, 23]. However, using the same Simoa detecting method, a Chinese group did not find any correlation of NfL with episodic memory, information processing speed, executive function, or visuospatial function after adjusting for age, gender, and education. In our results, the plasma NfL had a significant correlation after adjustment for age and other covariates, not only with global cognition, but also with the other four cognitive domains. This suggested that, although NfL was regarded as a non-specific biomarker for neurological diseases [12], it still has value in the monitoring of AD cognitive deterioration. However, many correlations could not be observed when we classified participants into three groups according to CDR levels of 0, 0.5, and ≥1. The paradoxical results may reflect the underlying diverse pathophysiological conditions in different cognitive status [46].

The traditional amyloid cascade theory emphasized that Aβ as an initial factor triggers the following Tau pathology [1]. However, the plasma amyloid biomarkers (Aβ_1-42_, Aβ_1-40_, Aβ_1-42_/Aβ_1-40_) in our study had a relatively weaker correlation only with MMSE, Memory, and Attention. Previous CSF and PET studies [47, 48] indicated that, the amyloid-related biomarkers reflected the earliest pathological change but tended to reach a plateau as the disease progressed to the dementia stage. Since the plasma Aβ level was supposed to reflect the central nervous change [49], blood Aβ level may also be saturated in the symptomatic individuals. However, in the CDR subgroups, the trend is ambiguous and inconsistent, probably because of the small sample size. Further large-sampled longitudinal studies need to be conducted to demonstrate the dynamic of plasma Aβ levels along the clinical spectrum of AD. A previous experimental study showed that neurons exposed to Aβ had increased synthesis and secretion of tau [50]. These neurons may eventually degenerate and develop tangle pathology, which may also drive the increase of p-tau in the blood. Therefore, regarding AD as a whole disease continuum, plasma p-tau181, as a more sensitive and clinical-relevant blood-based biomarker, may be superior to Aβ.

## Limitations

There are several limitations in our study. Firstly, the biomarkers in this study were only detected once without longitudinal measurements. However, we separated the participants into six groups according to CDR scores and cognitive diagnosis to simulate the AD spectrum. Future prospective studies are needed to verify our findings. Secondly, the participants in our studies were from two different resources with unavoidable imparity. However, there was no significant difference in some dementia-related risk factors, such as smoking, alcohol consumption, hypertension, and diabetes mellitus (Table 1). Age, gender, education year, and APOE were adjusted in the multivariate statistical models. Thirdly, some neuropsychological tests might have “ceiling effects” (e.g. Go/No-Go Task) or “floor effects” (e.g. Auditory Verbal Learning Test), which may conceal the relationship between the biomarkers and the cognitive domains. Lastly, we diagnosed AD based on the clinical standard rather than pathological evidence of CSF or amyloid/tau PET. Lacking a golden standard impeded us from the classifying of “ATN” framework [51] or Receiver Operating Characteristic analysis.

## Conclusions

In conclusion, we found plasma p-tau181 increased along the clinical continuum of AD. Plasma p-tau181 had a stronger correlation with cognitive domains than other biomarkers, especially in late-stage AD. Our study suggests that the plasma p-tau181 could reflect the AD pathology in vivo and may be a promising blood-based biomarker in clinical settings. Longitudinal studies are needed to verify these findings and provide more evidence of the association between plasma p-tau181 and clinical cognitive manifestations.

## Data Availability

The datasets used and/or analyzed during the current study are available from the corresponding author on reasonable request.

## Abbreviations

AD: Alzheimer’s disease
aMCI: amnestic mild cognitive impairment
aMCI-m: amnestic mild cognitive impairment multi-domains
aMCI-s: amnestic mild cognitive impairment single-domain
ANOVA: Analysis of Variance
APOE: Apolipoprotein E
CDR: Clinical Dementia Rating
CSF: cerebrospinal fluid
EDTA: Ethylene Diamine Tetraacetic Acid
MCI: mild cognitive impairment
MMSE: Mini-mental State Examination
MoCA-B: Montreal Cognitive Assessment-Basic
NC: normal cognition
NfL: neurofilament protein light chain
PET: positron emission tomography
p-tau181: phosphorylated tau 181
SAS: Shanghai Aging Study
SD: standard deviation
Simoa: Single molecule array
TMT: Trail Making Test
t-tau: total tau

## Acknowledgements

Not applicable

## Authors’ contributions

QZ, HX, and DD developed the original idea. ZX and XW searched the literature. ZX, WW, XL, SD, and LZ collected samples and data. XW and JY measured the blood biomarkers with support from HG. ZX and JL analyzed data. ZX and XW wrote the manuscript. QZ, HX, and DD revised the manuscript. All authors read and approved the final manuscript.

## Ethics approval and consent to participate

This study was approved by the Medical Ethics Committee of Huashan Hospital, Fudan University, Shanghai, China (No. 2009-195 and 2011-288). All participants and/or their legal proxy gave written informed consent to participate.

## Consent for publication

Not applicable

## Author details

^1^Institute of Neurology, Huashan Hospital, Fudan University, Shanghai, China; ^2^National Clinical Research Center for Aging and Medicine, Huashan Hospital, Fudan University, Shanghai, China; ^3^School of Biomedical Engineering, Shanghai Jiao Tong University, Shanghai, China; ^4^Department of Biostatistics, School of Public Health, Fudan University, Shanghai, China; ^5^Key Lab of Public Health Safety of the Ministry of Education, Fudan University, Shanghai, China.

