## Supplementary Table 1 for "Plasma biomarker profiles and the correlation with cognitive function across the clinical spectrum of Alzheimer’s disease"

Supplementary Table 1. Domain-specific cognition extracted from neuropsychological tests in the current study.

| Global cognition | Memory | Attention | Visuospatial function | Language | Executive function |
| --- | --- | --- | --- | --- | --- |
| MMSE | 'delayed recall'  in MMSE | 'registration'  in MMSE | 'overlapping imaging'  in MoCA-B | 'naming'  in MMSE | 'trail making' & 'similarity'  in MoCA-B |
|  | 'delayed recall'  in MoCA-B | 'attention'  in MoCA-B | 'copy'  in Rey-Osterrieth Complex Figure | 'naming'  in MoCA-B | 'part B completion time'*  in Trail Making Test (TMT) |
|  | 'delayed recall'  in Auditory Verbal Learning Test | 'inhibitory control'  in Conflicting Instructions Task (Go/No-Go Task) | 'copy'  in Stick Test | Boston Naming Test |  |
|  |  | Symbol Digit Modalities Test |  | 'objects naming'  in Modified Common Objects Sorting |  |

Note: The score of each test was transformed into accuracy (acquired score/total score, %) in all participants [1].

*The performance time values of TMT were divided into three categories to calculate the accuracy according to previously published normative data [2]. The TMT time more than mean + 1.5 standard deviation (SD) was regarded as poor executive function and was assigned 0 score. The TMT time less than mean – 1.5 SD was regarded as good executive function and was assigned 2 scores. Other participants were assigned 1 score. Then, the new TMT scores were also converted to accuracy and *Z*-scores.

Abbreviations: MMSE, mini-mental state examination; MoCA-B, Montreal Cognitive Assessment-Basic; TMT, Trail Making Test.
